# COVID-19: Predictive Mathematical Models for the Number of Deaths in South Korea, Italy, Spain, France, UK, Germany, and USA

**DOI:** 10.1101/2020.05.08.20095489

**Authors:** Athanasios S. Fokas, Nikolaos Dikaios, George A. Kastis

## Abstract

We have recently introduced two novel mathematical models for characterizing the dynamics of the cumulative number of individuals in a given country reported to be infected with COVID-19. Here we show that these models can also be used for determining the time-evolution of the associated number of deaths. In particular, using data up to around the time that the rate of deaths reaches a maximum, these models provide estimates for the time that a plateau will be reached signifying that the epidemic is approaching its end, as well as for the cumulative number of deaths at that time. The plateau is defined to occur when the rate of deaths is 5% of the maximum rate. Results are presented for South Korea, Italy, Spain, France, UK, Germany, and USA. The number of COVID-19 deaths in other counties can be analyzed similarly.

## Introduction

COVID-19 is the third coronavirus to appear in the human population in the past two decades; the first was the severe acute respiratory syndrome coronavirus SARS-CoV that caused the 2002 outbreak; the second was the Middle East syndrome coronavirus MERS-CoV responsible for the 2012 outbreak. COVID-19 has now caused a pandemic, which poses the most serious global public health threat since the devastating 1918 H1N1 influenza pandemic that killed approximately 50 million people (in proportion to today’s population, this corresponds to 200 million people). The unprecedented mobilization of the scientific community has already led to remarkable progress towards combating this threat, such as understanding significant features of the virus at the molecular level, see for example *(1)* and *(2)*. In addition, international efforts have intensified towards the development of specific pharmacological interventions; they include, clinical trials using old or relatively new medications and the employment of specific monoclonal antibodies, as well as novel approaches for the production of an effective vaccine. For example, there are ongoing linical trials testing the synthetic protein tocilizumab that binds inerleukin-6 (tocilizumab is often used in rheumatoid arthritis), as well as clinical trials involving the infusion to infected patients of plasma from individuals who have recovered from a SARS-CoV-2 infection *(3)*. Also, taking into consideration that the *complement pathway* is an integral component of the innate immune response to viruses, and that complement deposits are abundant in the lung biopsies from SARS-CoV-2 patients indicating that this system is presumably overacting *(6)*, it has been suggested that anti-complement therapies may be beneficial to SARS-CoV-2 patients. Further support for this suggestion is provided by earlier studies showing that the activation of various components of the complement system exacerbates the acute respiratory distress syndrome disease associated with SARS-CoV *(6)*. The Federal Drug Administration of USA has granted a conditional approval to the anti-viral medication Remdesivir *(7)*. Unfortunately, the combination of the anti-viral medications lopinavir and ritonavir that are effective against the human immunodeficiency virus has not shown any benefits *(4)*. Similarly, the combination of the anti-malarial medication hydroxychloroquine and the antibiotic azithromycin, is not only ineffective but can be harmful *(5)*.

The scientific community is also playing an important role in advising policy makers of possible non-pharmacological approaches to limit the catastrophic impact of the pandemic. For example, following the analysis in *(8)* of two possible strategies, called mitigation and suppression, for combating the epidemic, UK switched from mitigation to suppression. Within this context, in order to design a long-term strategy, it is necessary to be able to predict important features of the COVID-19 epidemic, such as the final accumulative number of deaths. Clearly, this requires the development of predictive mathematical models.

In a recent paper *(9)* we presented a model for the dynamics of the accumulative number of individuals in a given country that are reported at time *t* to be infected by COVID-19. This model is based on a particular ordinary differential equation of the Riccati type, which is specified by a *constant parameter* denoting the final total number of infected individuals, and by a *time-dependent function;* this function and the above parameter capture the effect of the basic characteristics of the COVID-19 infection as well the effect of the various measures taken by a given country for combating the spread of the infection. Remarkably, although the above Riccati equation is a nonlinear equation depending on time-dependent coefficients, it can be solved in closed form. Its solution depends on the above parameter and function, as well as on a parameter related to the constant arising in integrating this equation.

In the particular case that the associate time-dependent function is a constant, the explicit solution of the above Riccati equation becomes the classical *logistic formula*. It was shown in (9) that although this formula provides an adequate fit of the given data, is *not* sufficiently accurate for predictive purposes. In order to provide more accurate predictions, we introduced two novel models, called *rational* and *birational*.

Here we will show that the Ricatti equation introduced in *(9)* can also be used for determining the time evolution of the number, *N(t)*, of deaths in a given country caused by the COVID-19 epidemic. This Riccati equation is specified by the parameter *N_f_*, denoting the final total number of deaths, and by the function *α(t)*; this function together with *N_f_* model the effect of the basic characteristics of the SARS-CoV-2 infection as well as the cumulative effect of the variety of different measures taken by the given country for the prevention and the treatment of this viral infection. In the case that *α(t)* is a constant, denoted by *k*, the solution of the above Riccati equation becomes the well-known logistic formula,

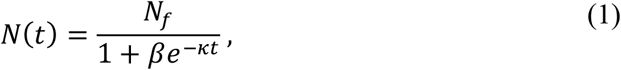

where *β* is the constant of integration. In the case that *α(t)* is the rational function *kd/1+kt*, the Riccati equation gives the rational formula; this is obtained from the logistic formula by simply replacing the exponential function with the function *(1+dt)^-k^*, where d and k are constants. The birational formula is similar with the rational formula, but the associated parameters are different before and after a fixed time X; this constant parameter is ether *T* or in the neighborhood of *T*, where *T denotes the time that the rate of deaths reaches a maximum*. The point on the curve depicting *N* as a function of *t* corresponding to *t=T* is known as the *inflection point*.

It turns out that the birational formula, in general, yields better predictions that the rational, which in turn provides better predictions than the logistic. Also, in general, the birational curve is above the curve obtained from the data, whereas the rational curve is below. Thus, the *birational and rational formulas may provide upper and lower bound estimates for the number of deaths at the plateau*. The birational formula can be employed only after the curve approaches its sigmoidal part (a precise criterion of when the birational can be used is discussed in the methods section). For this reason, at the present time the birational formula cannot be used for predictions applicable to the epidemics of Germany and USA.

Fig. 1 depicts the total number of deaths as a function of time after the day that 25 days had occurred, for the epidemics in South Korea, Italy, Spain, France, UK, Germany, and USA.

**Fig. 1.**
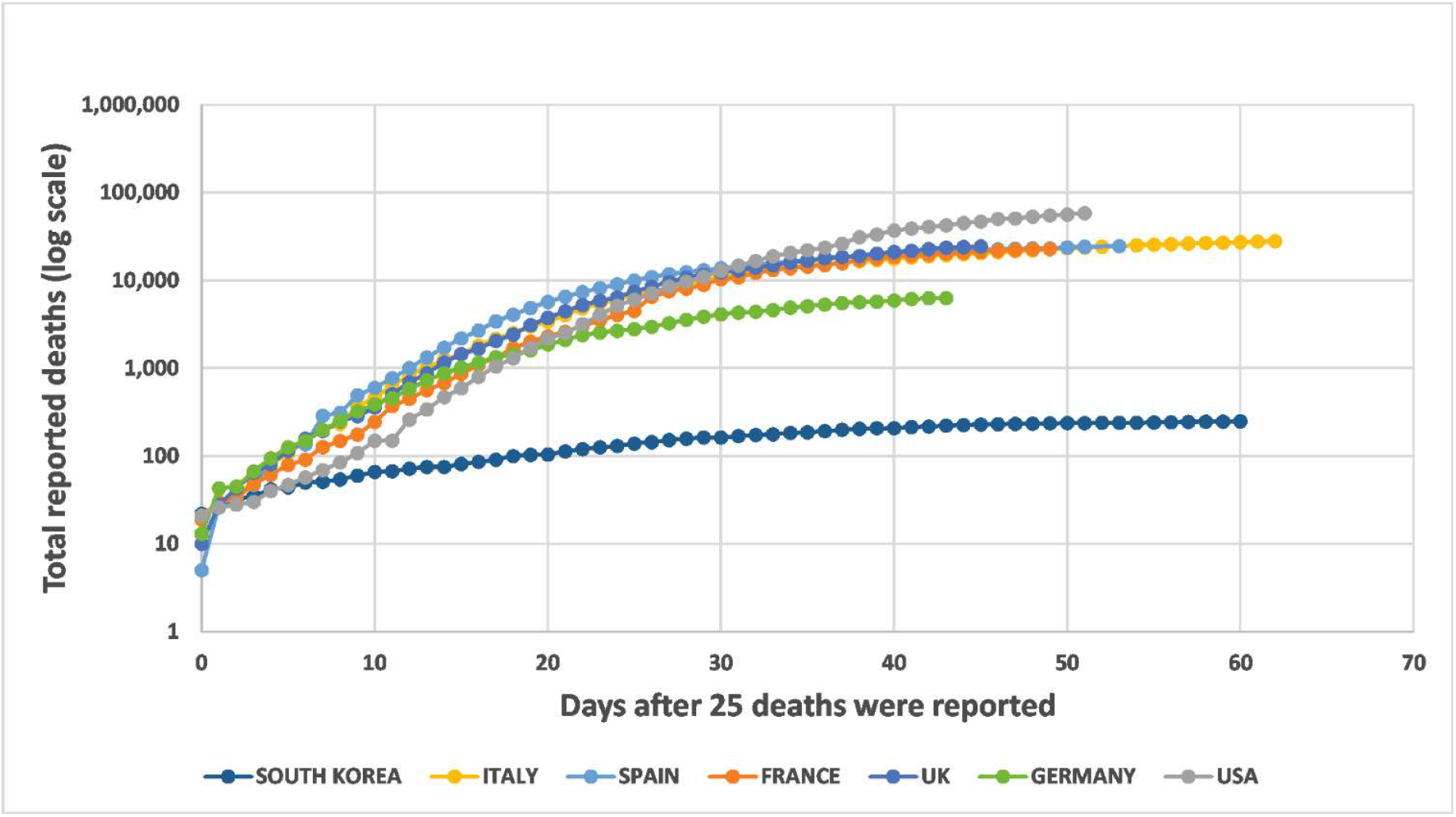
Total deaths vs days after 25 deaths were reported. COVID-19 virus epidenics in South Korea, Italy, Spain, France, United Kingdom, Germany and United States of America: Total cumulative number of deaths due to SARS-CoV-2 reported up to May 1, 2020, as a function of days after the day that 25 deaths were reported.

In this work we implemented the following tasks: (i) We determined the parameters specifying the logistic, rational, and birational formulas using the above data until May 1,2020. In this way it becomes evident that each of these models *can* fit the data. (ii) We also determined the parameters of the above models using data only a subset of the data. Then, by comparing the predictions of the resulting formulas with the remaining actual data it becomes clear that the rational and birational models generate more accurate predictions. Furthermore, for the cases of Italy and Spain the birational formula is above the data points, suggesting that the birational model may provide an upper bound (as noted earlier, for the epidemics of Germany and USA we fit only the logistic and the rational models). (iii) We computed the time of the plateau as well as the value of *N* at the plateau using the logistic and the rational models, as well as the birational model in the cases that it is applicable. *The plateau is defined as the point when the rate of deaths is 5% of the maximum rate*.

By implementing the above tasks we find the following estimates for the dates that the plateau will be reached as well as for the number of deaths when this occurs: South Korea plateau on May 18, 2020 with 258 deaths; Italy plateau on June 19, 2020 with 34,399 deaths; Spain plateau on May 25, 2020 with 27,090; France on May 13, 2020 with 26,619 deaths; UK on June 25, 2020 with 38,487 deaths; Germany on June 16, 2020 with 8,702 deaths; USA on June 1, 2020 with 78,532 deaths. The estimates for Italy and Spain may be upper bounds, whereas the remaining estimates are lower bounds.

## Results

Table 1 presents the model parameters and the plateau characteristics for the three different models as applied to South Korea, Italy, Spain, France, and UK. For the epidemics of Germany and USA, only the parameters and characteristics of the logistic and rational models are presented; for these countries, the sigmoidal part of their curves had not been reached as of May 1, 2020, and hence the birational formula could not be used for accurate predictions. The calculation of the inflection point requires the time that the derivative of N becomes maximum. For computing this point we used the logistic model (the other models yield similar results). For South Korea, Italy, Spain, France, UK, Germany, and USA, the inflection point occurred at t=23, t=35, t=28, t=33, t=32, t=27, and t=39, respectively. This corresponds to March 25, April 4, April 6, April 11, April 15, April 15, and April 17, 2020, respectively.

**Table 1.**
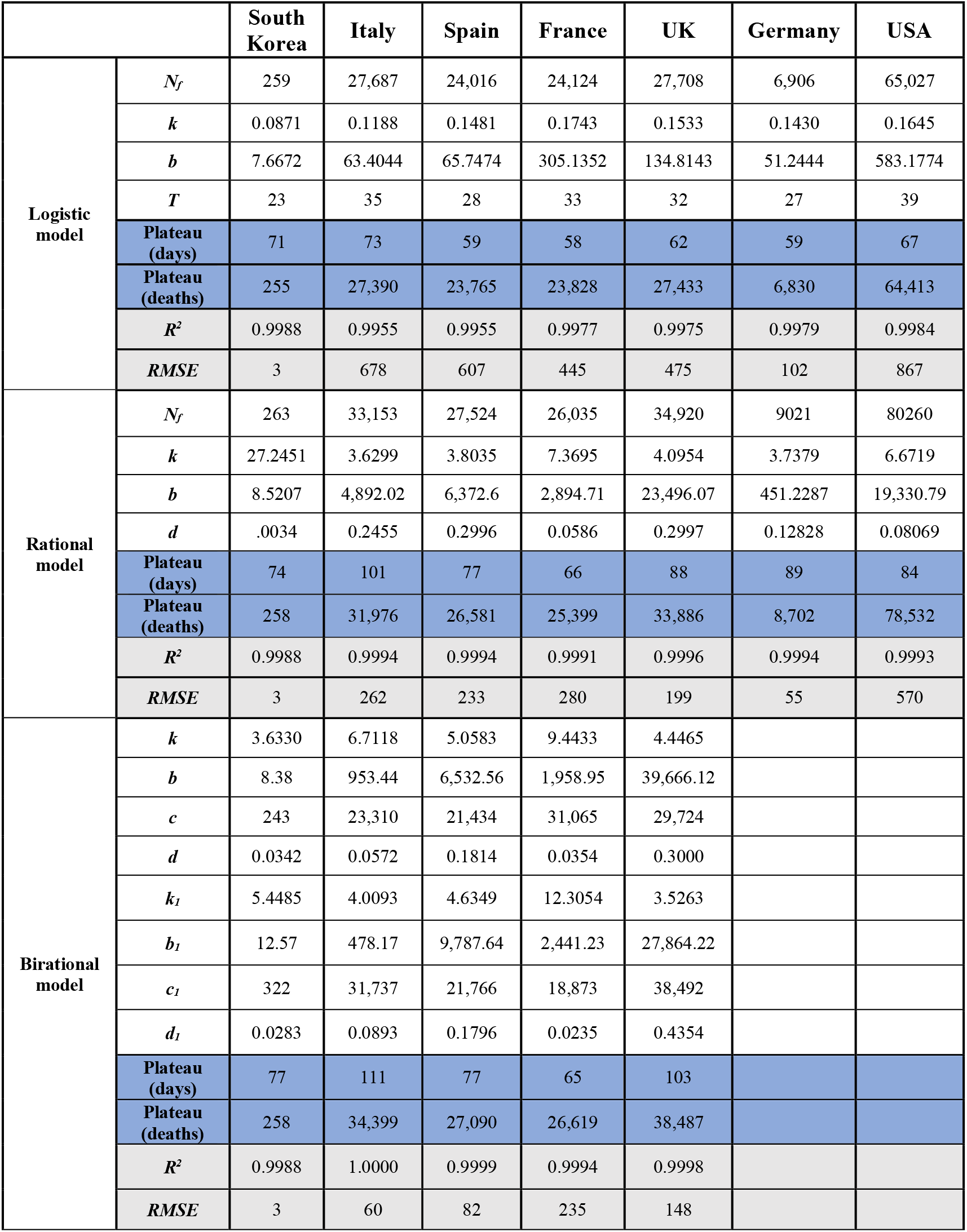
Model parameters and plateau characteristics. The model parameters and the plateau characteristics for the three different models are presented for South Korea, Italy, Spain, France, and UK. For the epidemics of Germany and USA only the parameters and characteristics of the logistic and rational models are presented. For those countries, the sigmoidal part of the curves had not been reached as of May 1, 2020 and h the birational formula could not be used for accurate predictions.

Fig. 2A presents the actual vs. predicted cumulative number of deaths, due to SARS-CoV-2, as a function of days after 25 deaths were reported, for the epidemic in Korea. A closer view of the fitting curves is presented in Fig. 2B. The logistic, rational, and birational formulas were trained with data up to May 1, 2020, which corresponds to T+37. The three models provide accurate fits, as quantified by the coefficient of determination, R2, and the Root-Mean-Square Error (RMSE). The logistic model predicts the plateau on May 12, 2020 (71 days after the day that 25 deaths were reported) with a total number of 255 deaths; the rational model predicts a plateau on May 15, 2020 (day 74) with 258 deaths; and the birational model predicts a plateau on May 18, 2020 (day 77) with 258 deaths.

**Fig. 2.**
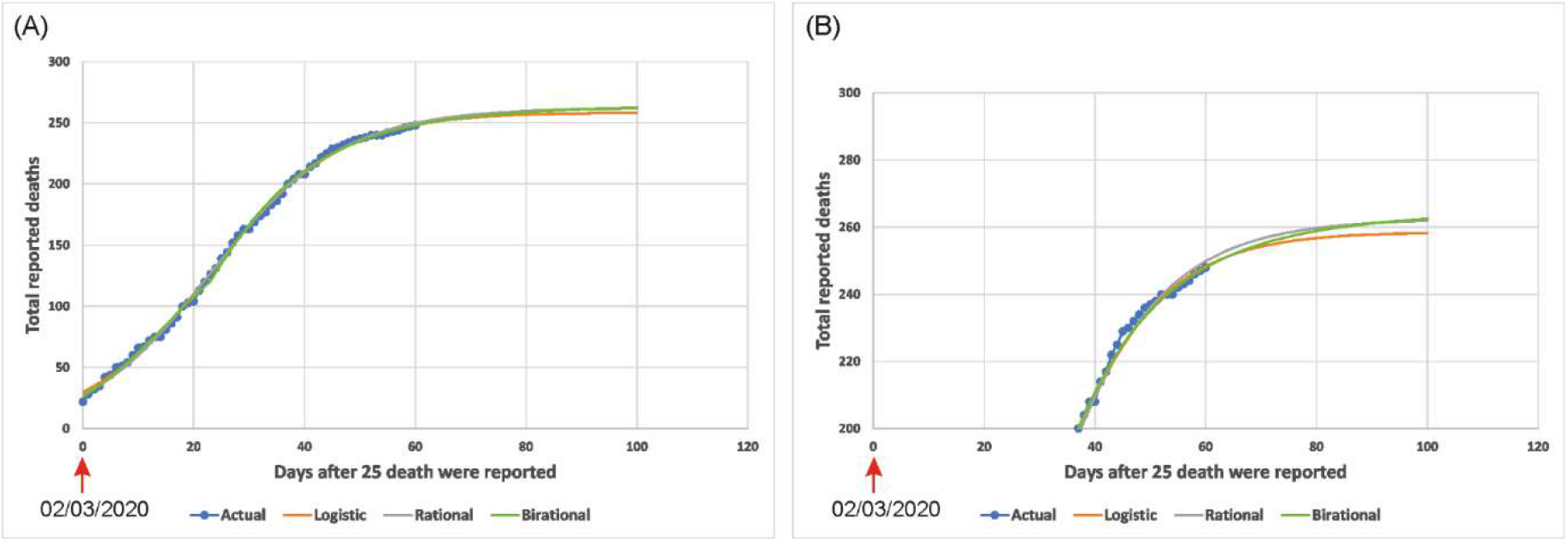
Actual vs Predicted SARS-CoV-2 deaths for South Korea. The Actual vs the Predicted cumulative number of reported deaths, due to SARS-CoV-2, are presented in **(A)** as a function of days after 25 deaths were reported for South Korea. A closer view of the fitted data is presented in **(B)**. The three models were trained using data up to May 1, 2020 (t=60). The inflection point occurred at t=23 which corresponds to March 25, 2020.

Fig. 3 presents the actual vs. predicted cumulative number of deaths, due to SARS-CoV-2, as a function of days after 25 deaths were reported, for Italy, Spain, France, UK, Germany, and USA. The three models were trained with data up to May 1, 2020, which corresponds to T+27, T+25, T+20, T+16, T+16, and T+14, respectively. For the epidemic of Italy (Fig. 3A), the logistic model predicts a plateau on May 12, 2020 (73 days after the day that 25 deaths were reported) with 27,390 deaths; the rational model predicts a plateau on June 9, 2020 (day 101) with 31,976 deaths; and the birational model predicts a plateau on June 19, 2020 (day 111) with 34,399 deaths. Clearly, the logistic model already underestimates the actual plateau day and the number of reported deaths, since on May 1, 2020, (last day of acquired data for this study) the number of deaths for Italy had reached 27,967.

**Fig. 3.**
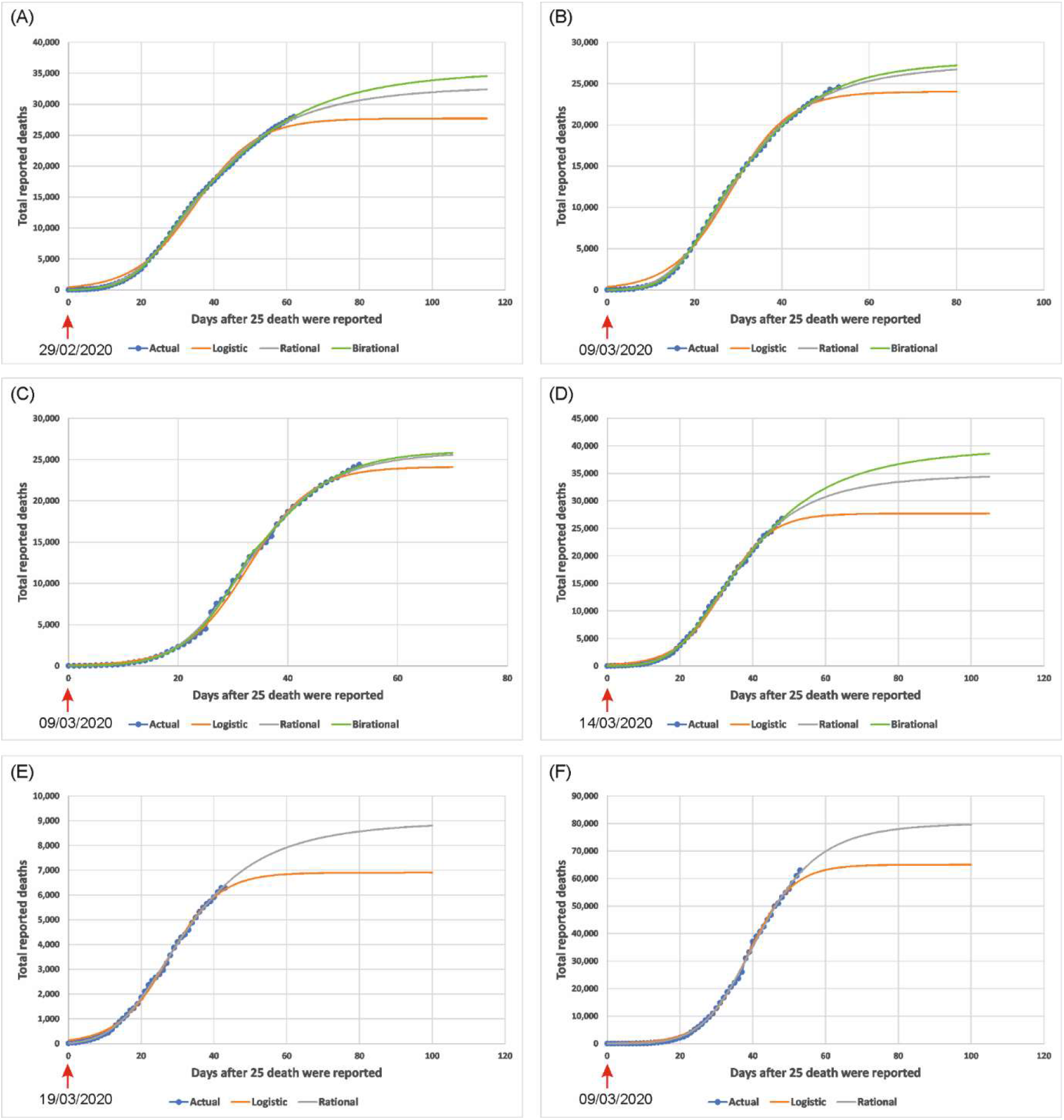
Actual vs Predicted SARS-CoV-2 deaths for various countries. The Actual vs the Predicted cumulative number of reported deaths, due to SARS-CoV-2, are presented as a function of days after 25 deaths were reported, for: **(A)** Italy, **(B)** Spain, **(C)** France, **(D)** UK, **(E)** Germany, **(F)** USA. The three models were trained with data up to May 1, 2020. This corresponds to t=62 data points for Italy, t=53 data points for Spain, t=53 data points for France, t=48 data points for UK, t=43 data points for Germany, and t=53 data points for USA. The inflection point occurs on April 4, 2020, for Italy; on April 6, 2020, for Spain; on April 11, 2020, for France; on April 15, 2020, for UK; on April 15, 2020, for Germany; and on April 17, 2020, for USA.

For the epidemic of Spain (Fig. 3B), the logistic model predicts a plateau on May 7, 2020 (day 59 after the day that 25 deaths were reported) with 23,795 deaths; the rational and birational models predicts a plateau on May 25, 2020 (day 77) with 26,581 and 27,090 deaths, respectively. Again, the logistic model already underestimates the actual plateau day and the number of reported deaths, since on May 1, 2020, the number of reported deaths for Spain had reached 24,543.

For the epidemic of France (Fig. 3C), the logistic model predicts a plateau on May 6, 2020 (58 days after the day that 25 deaths were reported) with 23,828 deaths; the rational model predicts a plateau on May 14, 2020 (day 66) with 25,399 deaths; and the birational model predicts a plateau on May 13, 2020 (day 65) with 26,619 deaths. The logistic model underestimates again the actual plateau day and the number of deaths, since on May 1, 2020, there were 24,376 confirmed deaths in France; thus, it is expected that the predicted number of the logistic model will be reached in a few days.

For the epidemic of the UK (Fig. 3D), the logistic model predicts a plateau on May 15, 2020 (62 days after the day that 25 deaths were reported) with 27,433 deaths; the rational model predicts a plateau on June 10, 2020 (day 88) with 33,886 deaths; and the birational model predicts a plateau on June 25, 2020 (day 103) with 38,487 deaths. The logistic model again underestimates the actual plateau day and the number of reported deaths, since on May 1, 2020, there were 26,771 confirmed deaths; it is expected that the predicted number of the logistic model will be reached in a few days.

For the epidemic of Germany (Fig. 3E), the logistic model predicts a plateau on May 17, 2020 (59 days after the day that 25 deaths were reported) with 6,830 deaths; the rational model predicts a plateau on June 16, 2020 (day 89) with 8,702 deaths. For the epidemic of the USA (Fig. 3F), the logistic model predicts a plateau on May 15, 2020 (67 days after the day that 25 deaths were reported) with 64,413 deaths; the rational model predicts a plateau on June 1, 2020 (day 84) with 78,532 reported deaths.

It is evident from the above that whereas each of the three equations (3), (10), and (11) can fit the data quite well, the predictive capacities of these formulas are *quite different*. This is further scrutinized in Fig. 4, where using the epidemic data from Italy, Spain, and France trained with data only until T+15, we compared the predictions of the resulting formulas with the actual data at later dates. For the trained data set, T+15 corresponds to t=50, t=43 and t=48, for Italy, Spain, and France, respectively. It is clear that each of the 3 models gives a different curve. Furthermore, for Italy (Fig. 4A) and Spain (Fig. 4B) the birational model provides an upper bound of the actual *N(t)*, whereas the rational model provides a lower bound which is better estimate than the one obtained via the logistic model. Regarding the birational model, for the cases of Italy and Spain the uppermost curves were obtained by using *X=T*. For France we experimented with different values of *X* for the birational model. For all such values, the birational curve was always above the rational curve; in order to obtain a curve that is above the actual *N(t)* for France (so that the resulting formula could perhaps yield an upper bound), it was necessary to choose *X=T-3*. Remarkably, the same result was obtained in (9) when examining an uppermost bound for the number of reported infected individuals.

**Fig. 4.**
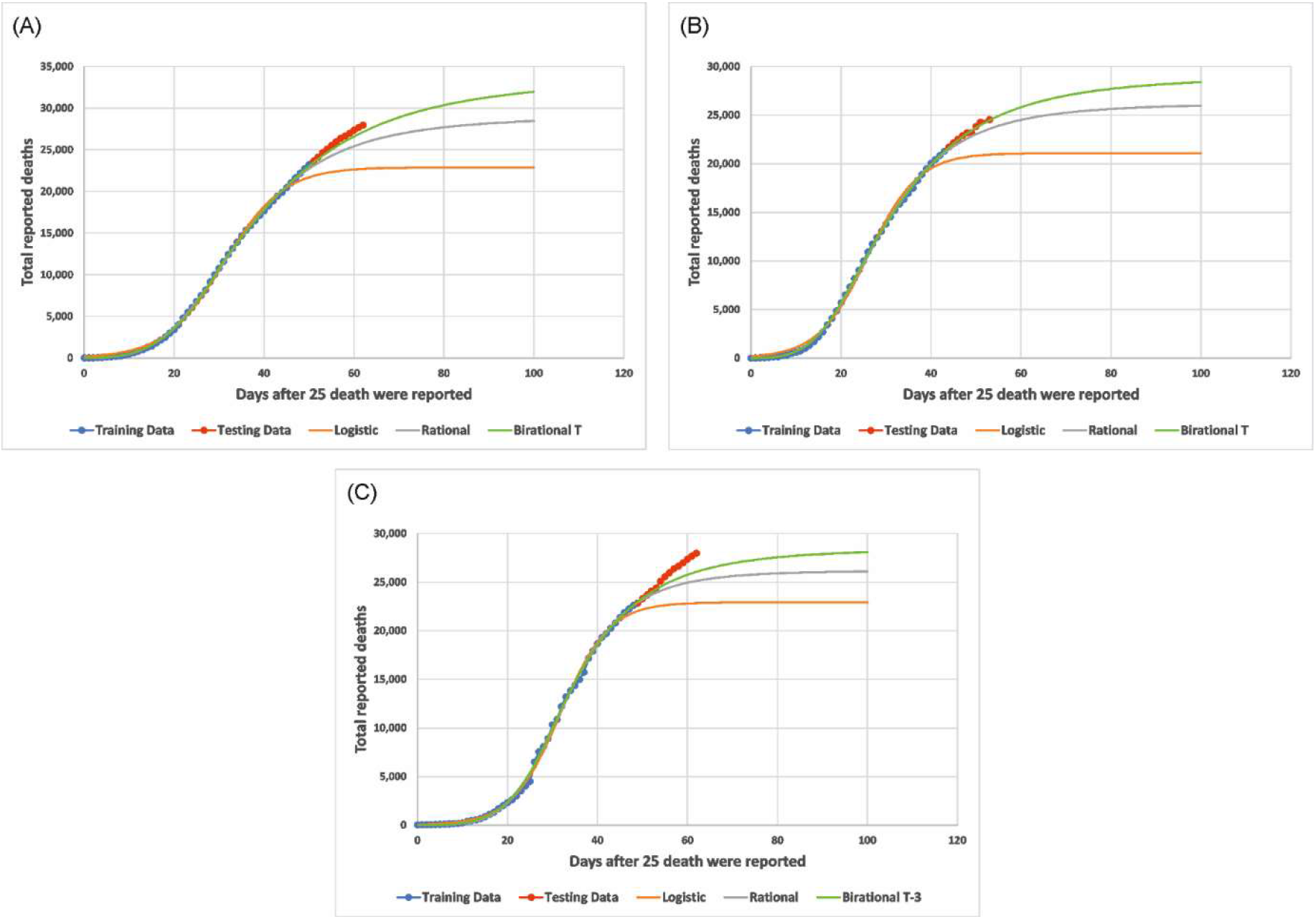
Predictions using a smaller training data set. Predictions using a smaller training data set for the cumulative number of reported deaths as a function of days after 25 deaths were reported for: **(A)** Italy, **(B)** Spain and **(C)** France. The prediction fits were obtained using training data up to T+15 for each country, which corresponds to t=50, t=43 and t=48, for Italy, Spain and France, respectively.

## Discussion

Several useful models elucidating aspect of the COVID-19 pandemic have already appeared in the literature, including *(10-19)*. These references represent only a tiny fraction of more than *3,000* publications that have appeared in the last four months in arXiv, medRxiv and bioRxiv.

Here, we have modelled the cumulative number *N* of deaths caused by SARS-CoV-2 in a given country as a function of time, in terms of a Riccati equation. This equation is specified by the constant *N_f_* and the function *α(t)*. Although this Riccati equation is a nonlinear ordinary differential equation containing time dependent coefficients, it can be solved in closed form. For appropriately chosen functions *α(t)*, the solution provides a flexible generalization of the classical logistic formula of equation (1) that has been employed in a great variety of applications, including the modelling of infectious processes.

The fact that *a* is now a function of *t* has important implications. In particular, it made it possible to construct the rational and birational models. All two models, as well as the logistic model provide good fits for the available data. However, as discussed in detail above, the rational and birational models provide more *accurate predictions*. Furthermore, *the birational model may provide an upper bound of the actual N(t), whereas the rational model yields a better lower bound that the logistic model*.

It is noted that our approach has the capacity for increasing continuously the accuracy of the predictions: as soon as the epidemic in a given country passes the time *T*, the rational model can be used; furthermore, when the sigmoidal part of the curve is approached, the rational model can be supplemented with the birational model. Also, as more data become available, the parameters of the rational and of the birational models can be re-evaluated; this will yield better predictions.

By formulating *N(t)* in terms of a Riccati equation, and by exploring the flexibility of the arbitrariness of *α(t)*, it is possible to decipher basic physiological mechanisms dictating the evolution of *N(t)*. In particular, following the transient stage of the epidemic, it may be envisioned that *α(t)* becomes a function of *N* instead of a function of *t*. This is indeed the case:by plotting *α* in terms of *N* is possible to scrutinize a posteriori their relationship; we find that after *t* approximately equal to *T-7* the relation between *α* and *N/25* becomes, remarkably, *linear*, see Fig. 5.

**Fig. 5.**
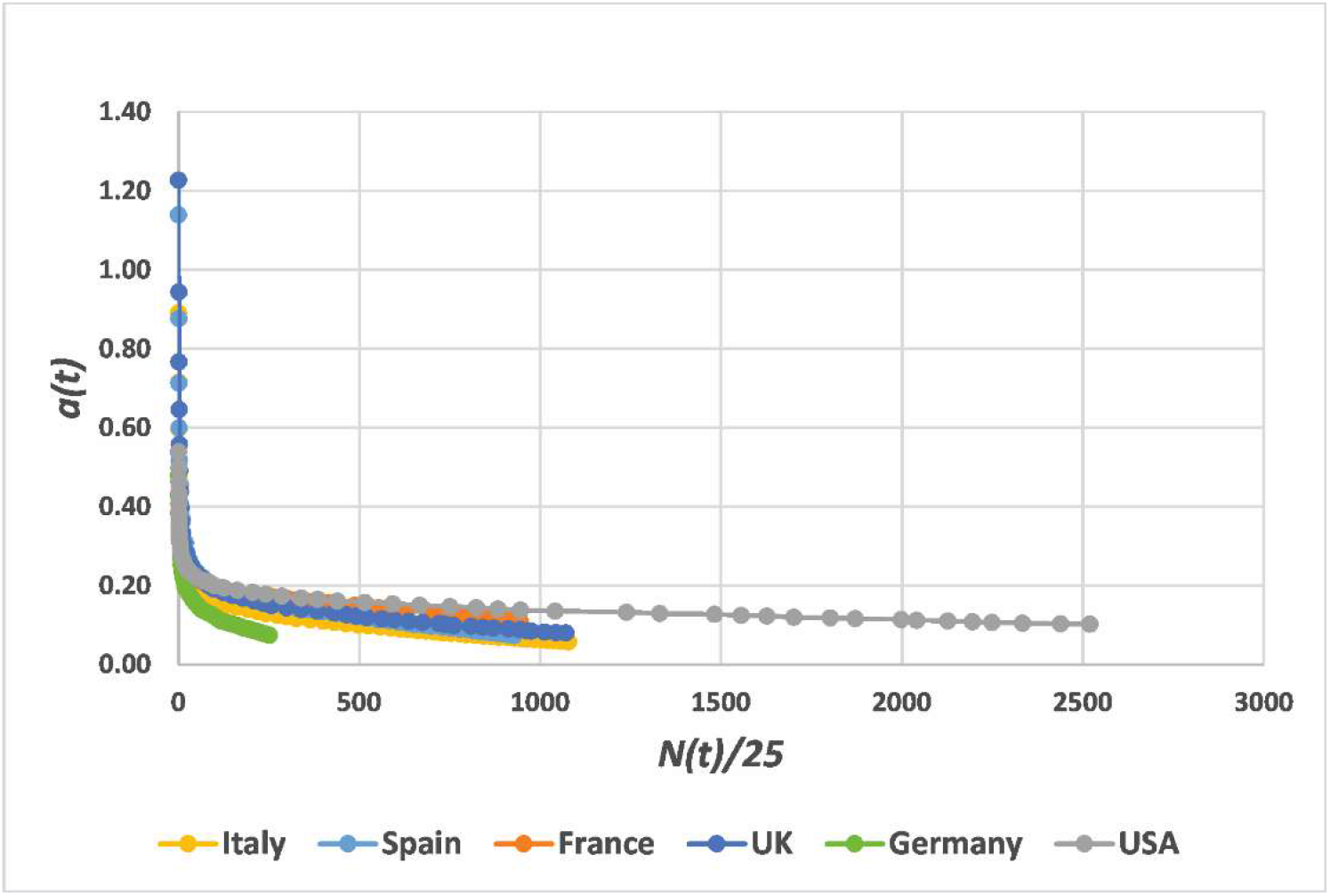
Plot of *a(t)* as a function of *N(t)/25* for the COVID-19 virus infection. The function *a(t)* is plotted as a function of *N(t)/25* for Italy, Spain, France, UK, Germany, and USA. For these countries, after *t* around T-7, *a(t)* becomes the *same linear function* of *N(t)*.

As a result of the justifiably draconian measures institutionalized in many countries, the number of the total persons reported infected by COVID-19, in China, South Korea, many European countries, USA, and several other countries, has now passed the inflection point. For the above countries, as shown here, the curve depicting the number of deaths as a function of time is also past the inflection point. Following this expected decline of the “first wave” of infections, it is now contemplated that the lockdown measures will be eased. This appears vital, not only for economical but also for health considerations. Indeed, the psychological impact on the population at large of the current situation is substantial (20); furthermore, it is expected to worsen, especially due to the effect of the post-traumatic disorder. A relatively safe ‘exit’ strategy avoiding a humanitarian disaster is suggested in (21). In any case, whatever strategy is followed, it is natural to expect that the number of reported infected individuals as well as the number of deaths will begin to grow. At this stage the predictions of the models proposed in (9) and here will not be valid. However, these works will still be valuable: they can be used to compute the additional number of reported infected individuals and deaths caused by easing the lockdown measures. Let us hope that a prudent exit strategy is adopted so that these numbers will not be staggering.

## Materials and Methods

We obtained the time-series data for the Coronavirus Disease (COVID-19) for South Korea, Italy, Spain, France, UK, Germany, and USA, from the official site of the European Centre for Disease Prevention and Control (22). We arranged the data in the form of deaths, N, over time measured in days, after the day that the number of deaths reached 25.

Throughout this paper, the unknown parameters were determined by employing the *simplex algorithm*. This is based on an iterative procedure that does not need information regarding the derivative of the function under consideration. The simplex algorithm is particularly effective for cases where the gradient of the likelihood functions is not easy to calculate. The algorithm creates a ‘random’ simplex of *n + 1* points, where *n* is the number of the model parameters that need to be estimated. The constrained variation of the simplex algorithm (23, 24) available in MATLAB® was used for all models; an L_1_*-norm* was employed in the likelihood function to improve robustness (25). Random parameter initializations were used to avoid local minima. The simplex algorithm was chosen because it performed better than the nonlinear least-squares curve fitting algorithms evaluated in this work, namely, the Levenberg-Marquardt (26) and the trust-region-reflective algorithms (27).

The stability of the fitting procedure was established by using the following simple criterion: different fitting attempts based on the use of a fixed number of data points, must yield curves which have the same form beyond the above fixed points. In this way, it was established that the rational formula could be employed provided that data were available until around the time T, whereas the birational formula could be used only for data available well beyond *T*.

The fitting accuracy of each model was evaluated by fitting the associated formula on *all* the available data in a specified set. The relevant parameters specifying the logistic, rational, and birational models are given on table 1.

We *assume* that the function *N(t)* satisfies the ordinary differential equation

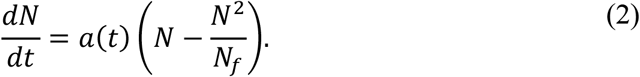

The general solution of this equation is given by (9)

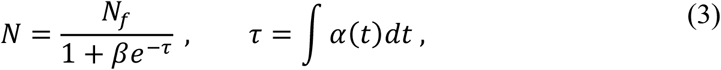

where *β* is the constant arising from integrating equation (2). In the particular case that *α(t)=k*, equation (3) becomes the logistic formula of equation (1) given in the introduction. If *α(t)* is given by the rational function below,

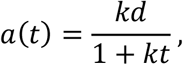

then equation (3) becomes the rational formula

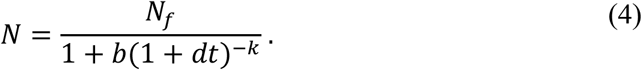

The birational formula is given by

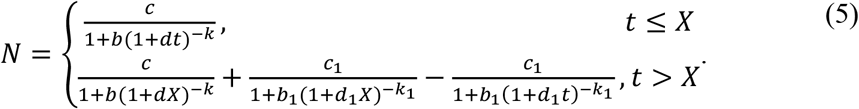

Where the fixed parameter *X* is either *T* or in the neighborhood of *T*. For the birational model

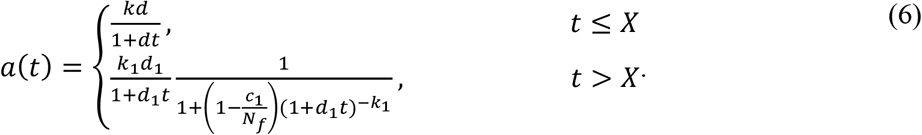

Letting in equation (5) *t* → ∞ we find:

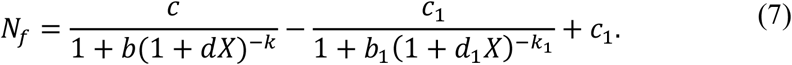

If *b, c, d, k*, are close to *b*_1_*, c*_1_*, d*_1_*, k*_1_, then *N_f_* is close to *c*_1_, and hence the value of *α(t)* after t=*X* is close to the value of *α(t)* before t= *X*.

The constant *T* can be computed by solving the equation obtained by equating to zero the second derivative of *N*. This implies that for the logistic and the rational models *T* is given respectively by

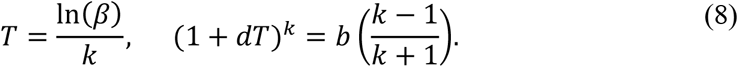

Similarly, for the birational model where the parameters *b, d, k*, in the second equations (8) are replaced with *b*_1_*, d*_1_*, k*_1_, respectively.

## Data Availability

We obtained the time-series data for the Coronavirus Disease (COVID-19) from the official site of the European Centre for Disease Prevention and Control.

https://www.ecdc.europa.eu/en/geographical-distribution-2019-ncov-cases

## Acknowledgments

General: A.S.F is grateful to several colleagues and in particular to the neurologist M. Dalakas and the mathematician S. Kourouklis for encouraging him to be involved with the modelling of COVID-19.

## Author contributions

A.S.F. conceived, designed and supervised the study. N.D. and G.A.K. performed the fitting experiments and analysis of the results. A.S.F. wrote the initial draft of the manuscript. All authors critically revised and improved the manuscript in various ways. All authors reviewed the manuscript and gave final approval for publication.

## Data and materials availability

All data needed to evaluate the conclusions in the paper are present in the paper and/or the Supplementary Materials.

